## Supplementary materials for "The Incidence and Prevalence of SARS-CoV-2 in the UK Population from the UKHSA Winter COVID Infection Study"

### Winter COVID-19 Infection Study Design

#### Cohort

The cohort sampled for this study was selected from previous participants of the COVID-19 Infection Study (CIS) [27], a household study. Participants were asked if they would consent to being contacted by UKHSA for future studies – those who selected yes were asked to enrol. Individuals eligible for the study lived in private households and were 3 years and older living in England or Scotland. Participants are asked a range of information about themselves and the results of SARS-CoV-2 diagnostic tests. The study was live from November 2023 to March 2024. Overall, there were 123,243 participants and 426,667 responses. Further information about how this data was collected can be found in the ONS quality and methodology information page for the study [28]. Information for the participants, including how to complete the questionnaire and conduct the tests are found on the ONS website [29]. The participant breakdowns by reference group are given in Supplementary Table (1).

#### Survey Design

The study had three core surveys, the first “participant” survey collected key identifying information used to link data, such as names, addresses and demographics. The second “main” survey asked further questions relating to individual characteristics (such as job, household size), current and new symptoms, key survey dates (including window start date, LFD taken date, symptom onset date) and lateral flow device (LFD) test results. The third “follow up” survey was given to those who reported a positive test in the “main” survey. The “follow up” survey asked participants to test on alternating days following their first positive test and submit the survey after 15 days.

For each survey participants responded via an online survey portal which guided them through each question. Participants were not offered financial incentives for the study but were given the lateral flow devices as part of the operations.

Participants were grouped into waves, and testing windows. Each participant received 14 tests at the start of the study and responded once to the main survey per wave, a total of 4 times over the course of the study. Each wave ran for approximately 4 weeks, with wave 1 running from 14 November to 14 December 2023, wave 2 from 12 December 2023 to 11 January 2024, wave 3 from 9 January to 8 February 2024 and wave 4 from 6 February to 7 March 2024. To allow for flexible timing in testing and response participants were given a window of 8 days to report their test. Participants were prompted at the start of their testing window and on day 5 to encourage response. Tests taken up to two days before the start of the testing window were allowed to be reported in the main survey.

#### Data Processing

Data were cleaned and duplicates dropped such that there was one record per participant per wave. Records were removed where linkage information was unavailable, or demographics information not given. This accounted for <1% of the participants and records. Samples were removed where the individual did complete the main survey but did not take an LFD test.

Where a participant did not know their symptom onset date they were prompted to give a placeholder date of the 15^th^ of the most recent month, for both the follow up and main survey. Therefore, values for symptom onset on these dates were assumed as missing. Demographic and participant characteristic information was captured in the first main survey response and not again unless there was a change in circumstance to avoid lengthy surveys. These demographic data were therefore filled forward. Participants who withdrew consent part way through or after the study were excluded from the analysis.

| **group** | **strata** | **n (participants)** | **%**  **(participants)** | **n**  **(responses)** | **% (responses)** | **%**  **(population)** |
| --- | --- | --- | --- | --- | --- | --- |
| all |  | 123243 |  | 426667 |  |  |
| age | 3 to 17 | 3886 | 3.15 | 11083 | 2.6 | 20.55 |
| age | 18 to 34 | 4220 | 3.42 | 11426 | 2.68 | 21.37 |
| age | 35 to 44 | 9959 | 8.08 | 29803 | 6.99 | 13.49 |
| age | 45 to 54 | 18448 | 14.97 | 61061 | 14.31 | 12.72 |
| age | 55 to 64 | 30387 | 24.66 | 108950 | 25.54 | 13.09 |
| age | 65 to 74 | 36813 | 29.87 | 133983 | 31.4 | 9.77 |
| age | 75 and over | 19530 | 15.85 | 70361 | 16.49 | 9 |
| sex | Female | 70189 | 56.95 | 241721 | 56.65 | 50.99 |
| sex | Male | 53054 | 43.05 | 184946 | 43.35 | 49.01 |
| location | North West | 13812 | 11.21 | 47568 | 11.15 | 12 |
| location | East Midlands | 8977 | 7.28 | 31266 | 7.33 | 7.86 |
| location | South East | 18782 | 15.24 | 65579 | 15.37 | 14.94 |
| location | London | 18118 | 14.7 | 61023 | 14.3 | 14.31 |
| location | South West | 12732 | 10.33 | 44551 | 10.44 | 9.19 |
| location | Scotland | 11469 | 9.31 | 39968 | 9.37 | 8.72 |
| location | East of England | 13969 | 11.33 | 48642 | 11.4 | 10.25 |
| location | Yorkshire and The Humber | 10634 | 8.63 | 36931 | 8.66 | 8.84 |
| location | West Midlands | 9722 | 7.89 | 33590 | 7.87 | 9.63 |
| location | North East | 5028 | 4.08 | 17549 | 4.11 | 4.27 |

Supplementary Table 1: The count (n) and proportion (%) of participants in each strata. The proportions are calculated as the count within a strata divided by the count within a give group.

### Additional results and plots

#### Prevalence

| Age group | Minimum prevalence (%) | Maximum prevalence (%) | Minimum prevalence date | Maximum prevalence date |
| --- | --- | --- | --- | --- |
| 3 to 17 years | 1.00  (95% CI: 0.608 to 1.60) | 4.34  (95% CI: 3.40 to 5.52) | 28/02/2024 | 23/12/2023 |
| 18 to 34 years | 0.725  (95% CI: 0.459 to 1.08) | 5.51  (95% CI: 4.51 to 6.62) | 02/03/2024 | 22/12/2023 |
| 35 to 44 years | 0.968  (95% CI: 0.696 to 1.31) | 5.98  (95% CI: 5.07 to 6.99) | 02/03/2024 | 22/12/2023 |
| 45 to 54 years | 0.767  (95% CI: 0.572 to 1.01) | 4.84  (95% CI: 4.14 to 5.62) | 29/02/2024 | 22/12/2023 |
| 55 to 64 years | 0.501  (95% CI: 0.370 to 0.662) | 4.07  (95% CI: 3.48 to 4.72) | 02/03/2024 | 22/12/2023 |
| 65 to 74 years | 0.401  (95% CI: 0.288 to 0.530) | 2.82  (95% CI: 2.41 to 3.29) | 02/03/2024 | 22/12/2023 |
| 75 years and over | 0.316  (95% CI: 0.216 to 0.440) | 2.52  (95% CI: 2.12 to 2.97) | 03/03/2024 | 22/12/2023 |

Supplementary Table 2: Age-stratified estimates of the date and value of the maximum and minimum prevalence.

| Location | Minimum prevalence (%) | Maximum prevalence (%) | Minimum prevalence date | Maximum prevalence date |
| --- | --- | --- | --- | --- |
| East Midlands | 0.759  (95% CI: 0.546 to 1.09) | 4.33  (95% CI: 3.60 to 5.12) | 01/03/2024 | 22/12/2023 |
| East of England | 0.751  (95% CI: 0.543 to 1.01) | 4.85  (95% CI: 4.12 to 5.70) | 01/03/2024 | 22/12/2023 |
| London | 0.746  (95% CI: 0.506 to 1.01) | 5.37  (95% CI: 4.60 to 6.27) | 01/03/2024 | 22/12/2023 |
| North East | 0.702  (95% CI: 0.492 to 1.07) | 3.92  (95% CI: 3.17 to 4.70) | 01/03/2024 | 22/12/2023 |
| North West | 0.645  (95% CI: 0.462 to 0.894) | 4.20  (95% CI: 3.53 to 4.91) | 02/03/2024 | 22/12/2023 |
| Scotland | 0.584  (95% CI: 0.408 to 0.801) | 3.92  (95% CI: 3.28 to 4.60) | 01/03/2024 | 22/12/2023 |
| South East | 0.768  (95% CI: 0.561 to 1.03) | 4.88  (95% CI: 4.18 to 5.71) | 01/03/2024 | 22/12/2023 |
| South West | 0.789  (95% CI: 0.587 to 1.08) | 4.72  (95% CI: 4.01 to 5.59) | 29/02/2024 | 22/12/2023 |
| West Midlands | 0.723  (95% CI: 0.520 to 1.02) | 4.17  (95% CI: 3.51 to 4.96) | 01/03/2024 | 22/12/2023 |
| Yorkshire and The Humber | 0.659  (95% CI: 0.471 to 0.908) | 3.98  (95% CI: 3.31 to 4.72) | 01/03/2024 | 22/12/2023 |

Supplementary Table 3: Location-stratified estimates of the date and value of the maximum and minimum prevalence.

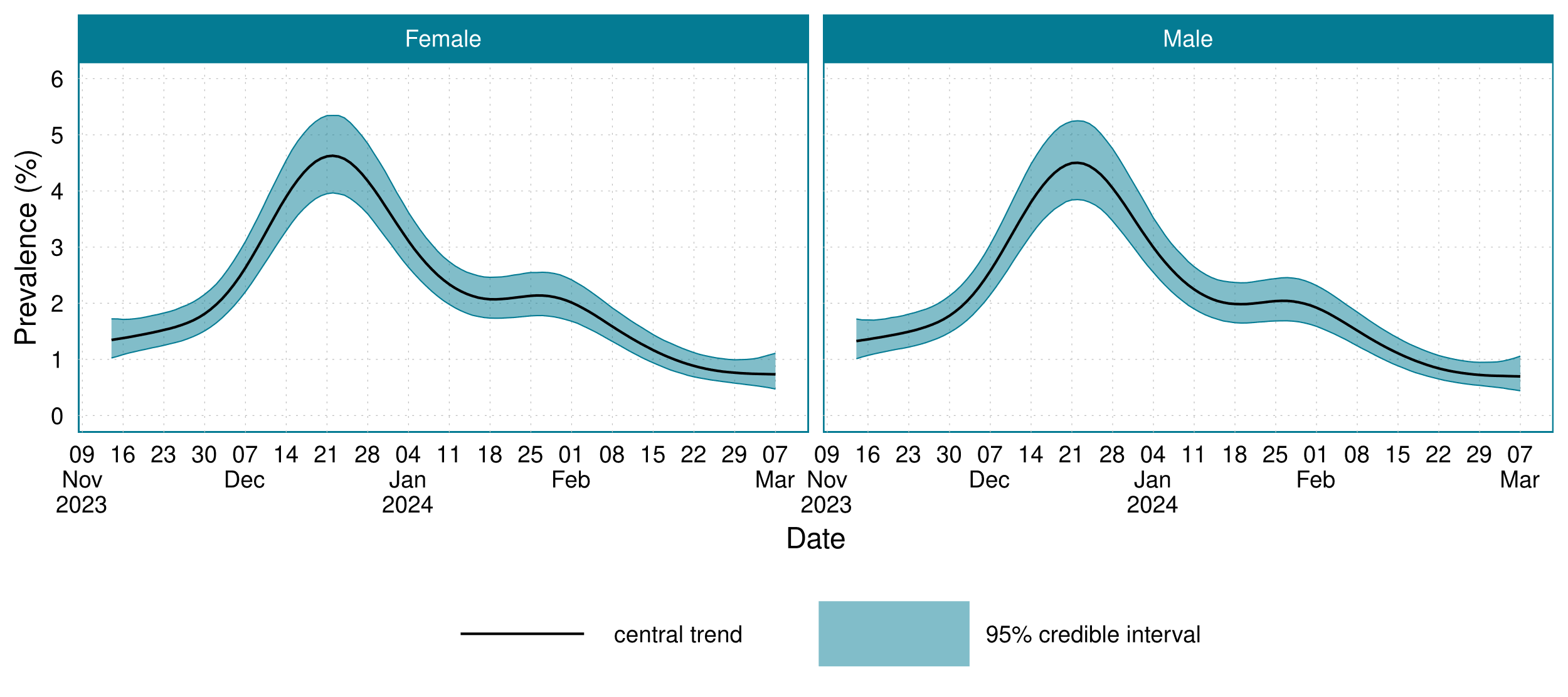

Supplementary Figure 1: The sex-stratified prevalence of SARS-CoV-2 in England and Scotland between 14^th^ November 2023 and 7^th^ March 2024.

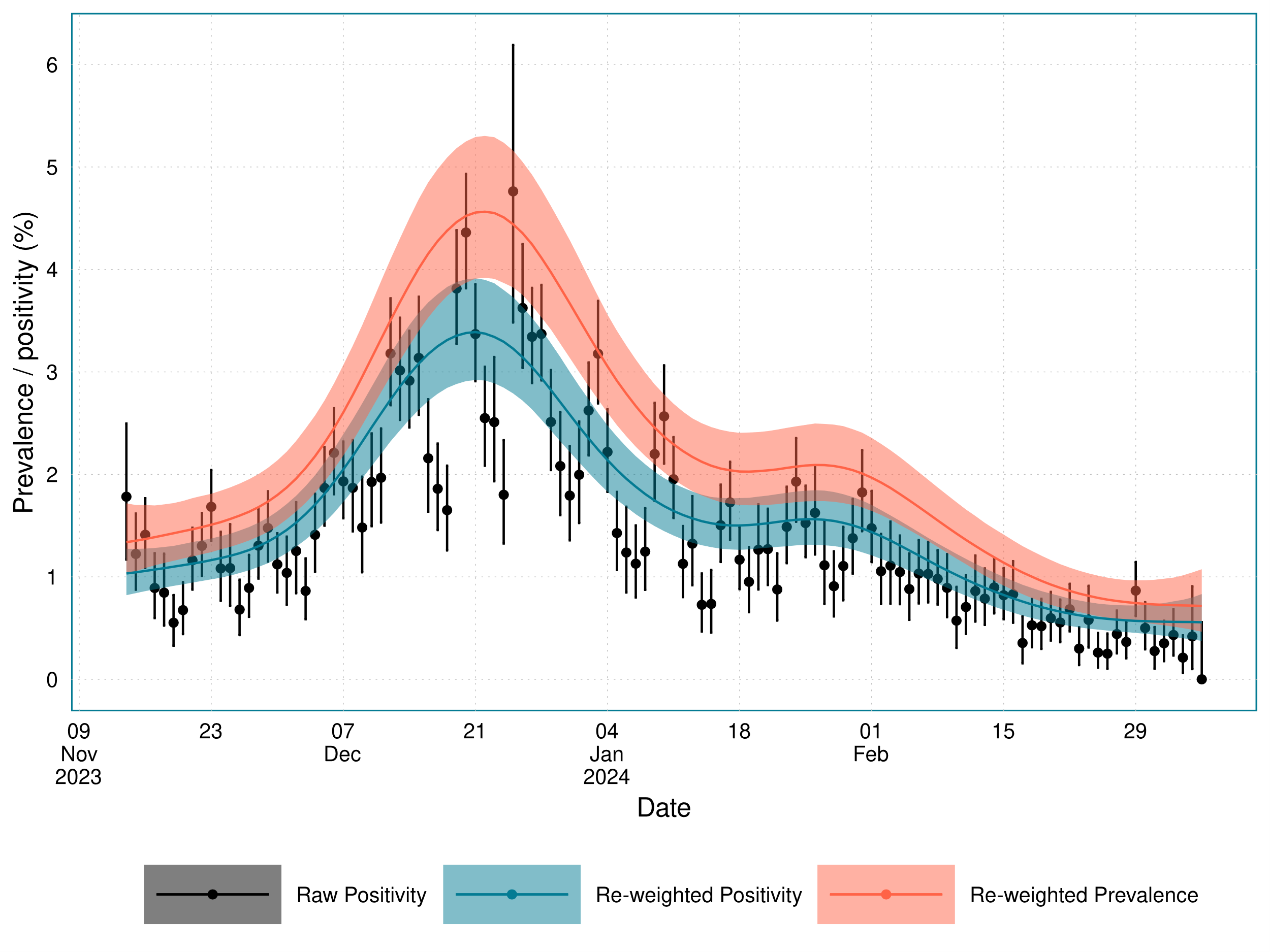

Supplementary Figure 2: Estimates of the poststratified positivity and prevalence, plotted alongside the daily sample positivity for the combined England and Scotland location.

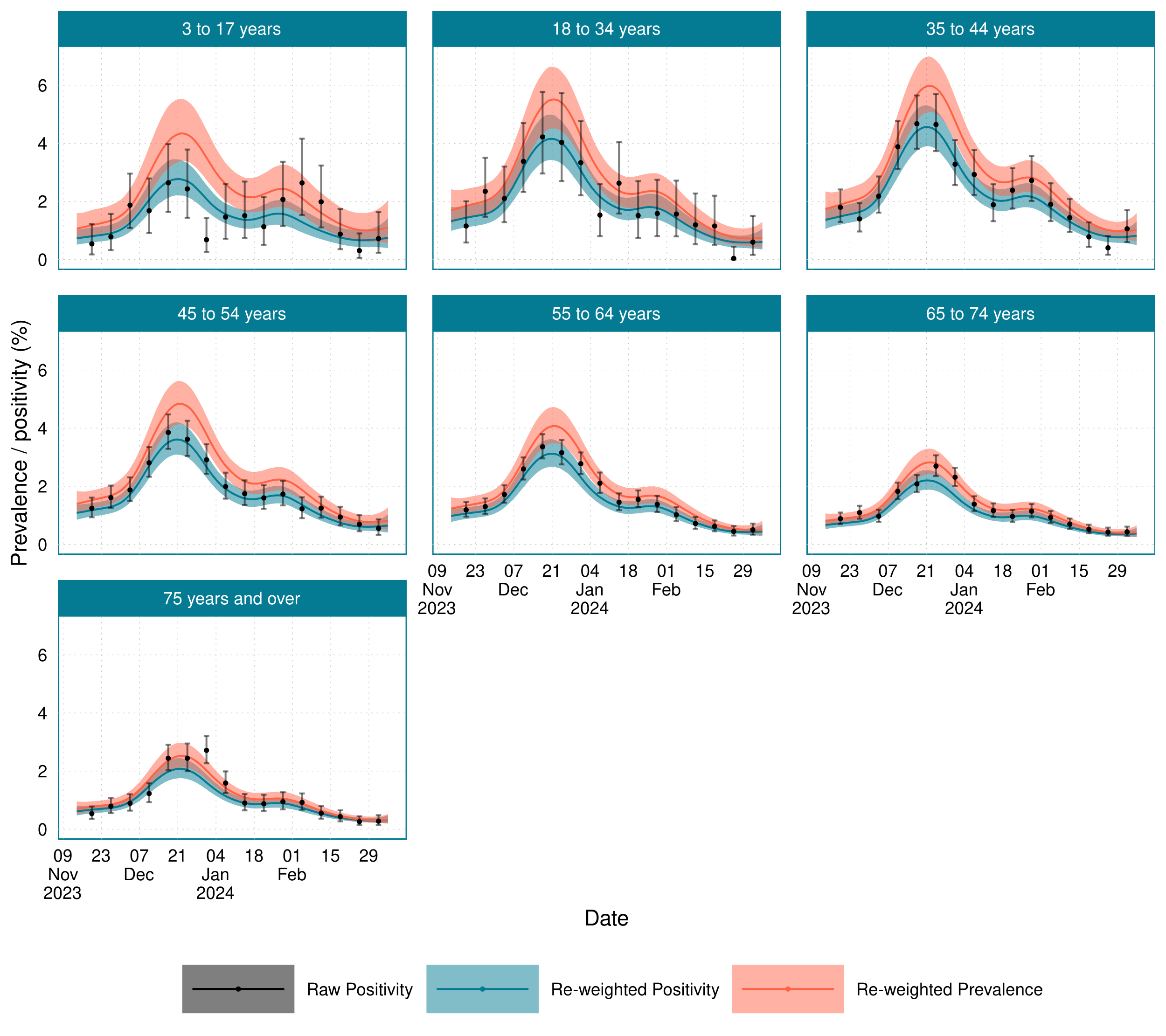

Supplementary Figure 3: Age-stratified estimates of the poststratified positivity and prevalence, plotted alongside the weekly sample positivity.

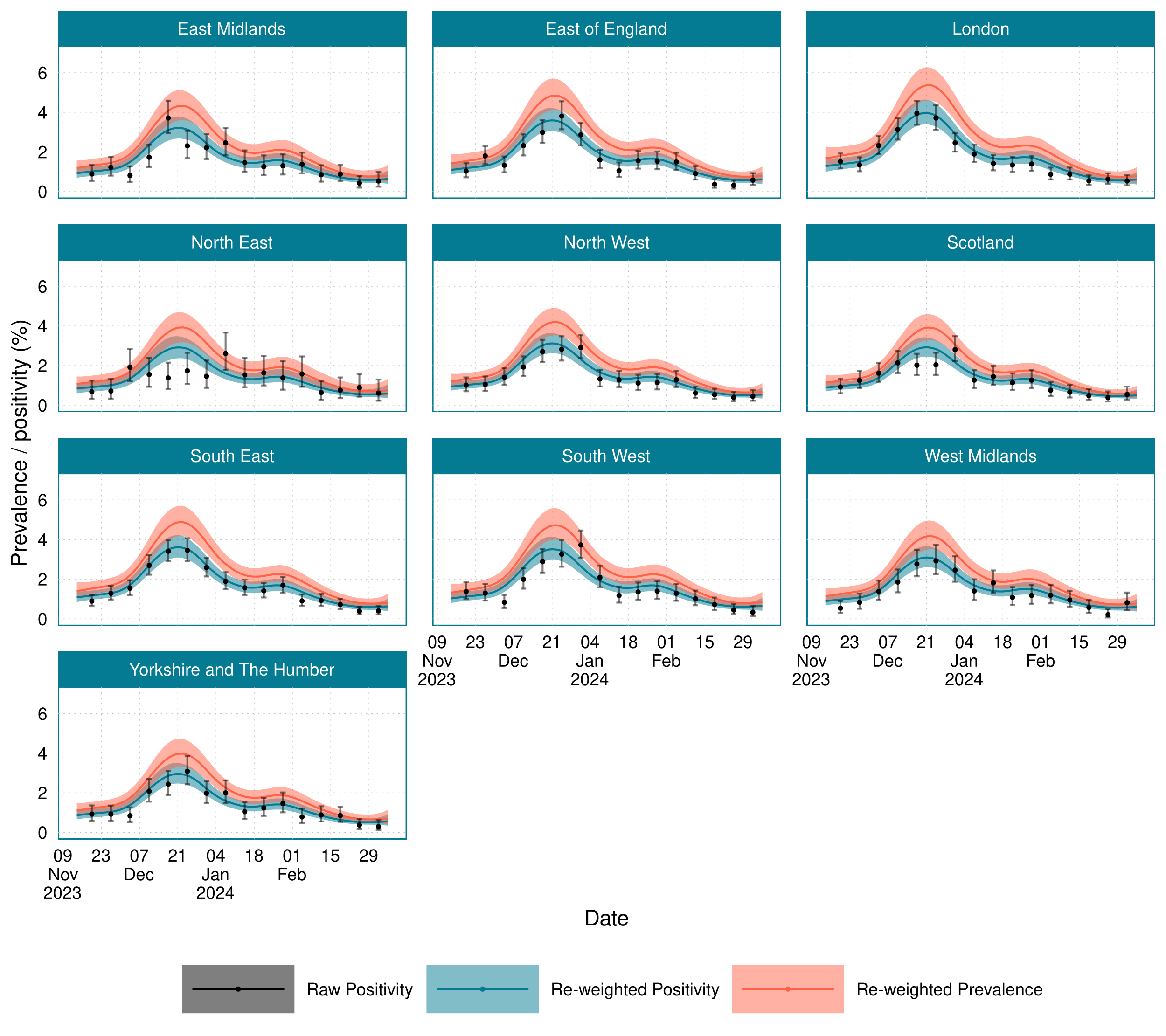

Supplementary Figure 4: Age-stratified estimates of the poststratified positivity and prevalence, plotted alongside the weekly sample positivity.

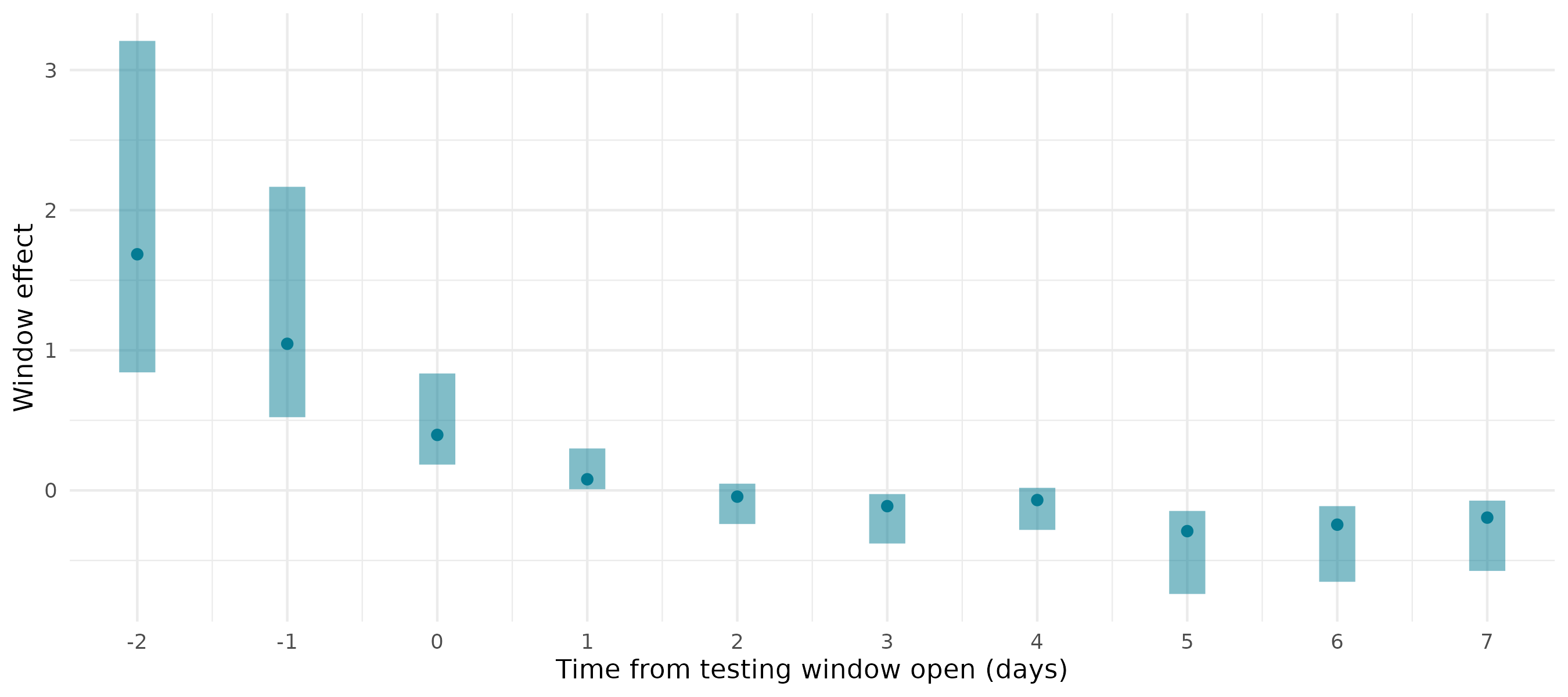

Supplementary Figure 5: Posterior estimates of the window effect $\beta_{i}^{\text{window}}$ across different days from window open

#### Incidence

| Age group | Minimum incidence  (new infections per 100,000 individuals per day) | Maximum incidence (new infections per 100,000 individuals per day) | Minimum incidence date | Maximum incidence date |
| --- | --- | --- | --- | --- |
| 3 to 17 years | 108  (95% CI: 64.2 to 178) | 504  (95% CI: 390 to 642) | 24/02/2024 | 18/12/2023 |
| 18 to 34 years | 75.3  (95% CI: 47.6 to 112) | 614  (95% CI: 502 to 754) | 25/02/2024 | 17/12/2023 |
| 35 to 44 years | 96.7  (95% CI: 69.3 to 133) | 643  (95% CI: 540 to 763) | 25/02/2024 | 17/12/2023 |
| 45 to 54 years | 78.9  (95% CI: 57.2 to 107) | 537  (95% CI: 453 to 631) | 25/02/2024 | 17/12/2023 |
| 55 to 64 years | 49.0  (95% CI: 36.1 to 64.9) | 432  (95% CI: 364 to 510) | 25/02/2024 | 17/12/2023 |
| 65 to 74 years | 38.1  (95% CI: 27.4 to 51.2) | 292  (95% CI: 247 to 347) | 25/02/2024 | 17/12/2023 |
| 75 years and over | 28.4  (95% CI: 19.6 to 39.4) | 249  (95% CI: 206 to 299) | 25/02/2024 | 17/12/2023 |

Supplementary Table 4: Age-stratified estimates of the date and value of peak incidence.

| Location | Minimum incidence  (new infections per 100,000 individuals per day) | Maximum incidence  (new infections per 100,000 individuals per day) | Minimum incidence date | Maximum incidence date |
| --- | --- | --- | --- | --- |
| East Midlands | 77.6  (95% CI: 54.6 to 112) | 474  (95% CI: 391 to 573) | 24/02/2024 | 17/12/2023 |
| East of England | 76.9  (95% CI: 54.3 to 105) | 531  (95% CI: 448 to 634) | 25/02/2024 | 17/12/2023 |
| London | 76.5  (95% CI: 51.6 to 105) | 592  (95% CI: 503 to 701) | 24/02/2024 | 17/12/2023 |
| North East | 71.6  (95% CI: 49.7 to 110) | 429  (95% CI: 344 to 523) | 24/02/2024 | 17/12/2023 |
| North West | 66.2  (95% CI: 46.8 to 90.6) | 460  (95% CI: 384 to 548) | 24/02/2024 | 17/12/2023 |
| Scotland | 59.6  (95% CI: 41.1 to 82.2) | 429  (95% CI: 356 to 510) | 24/02/2024 | 17/12/2023 |
| South East | 78.5  (95% CI: 56.8 to 107) | 535  (95% CI: 452 to 635) | 25/02/2024 | 17/12/2023 |
| South West | 80.4  (95% CI: 58.2 to 111) | 517  (95% CI: 433 to 618) | 25/02/2024 | 17/12/2023 |
| West Midlands | 73.9  (95% CI: 52.2 to 104) | 459  (95% CI: 379 to 553) | 24/02/2024 | 17/12/2023 |
| Yorkshire and The Humber | 67.5  (95% CI: 47.4 to 94.4) | 437  (95% CI: 361 to 526) | 25/02/2024 | 17/12/2023 |

Supplementary Table 5: Location-stratified estimates of the date and value of peak incidence

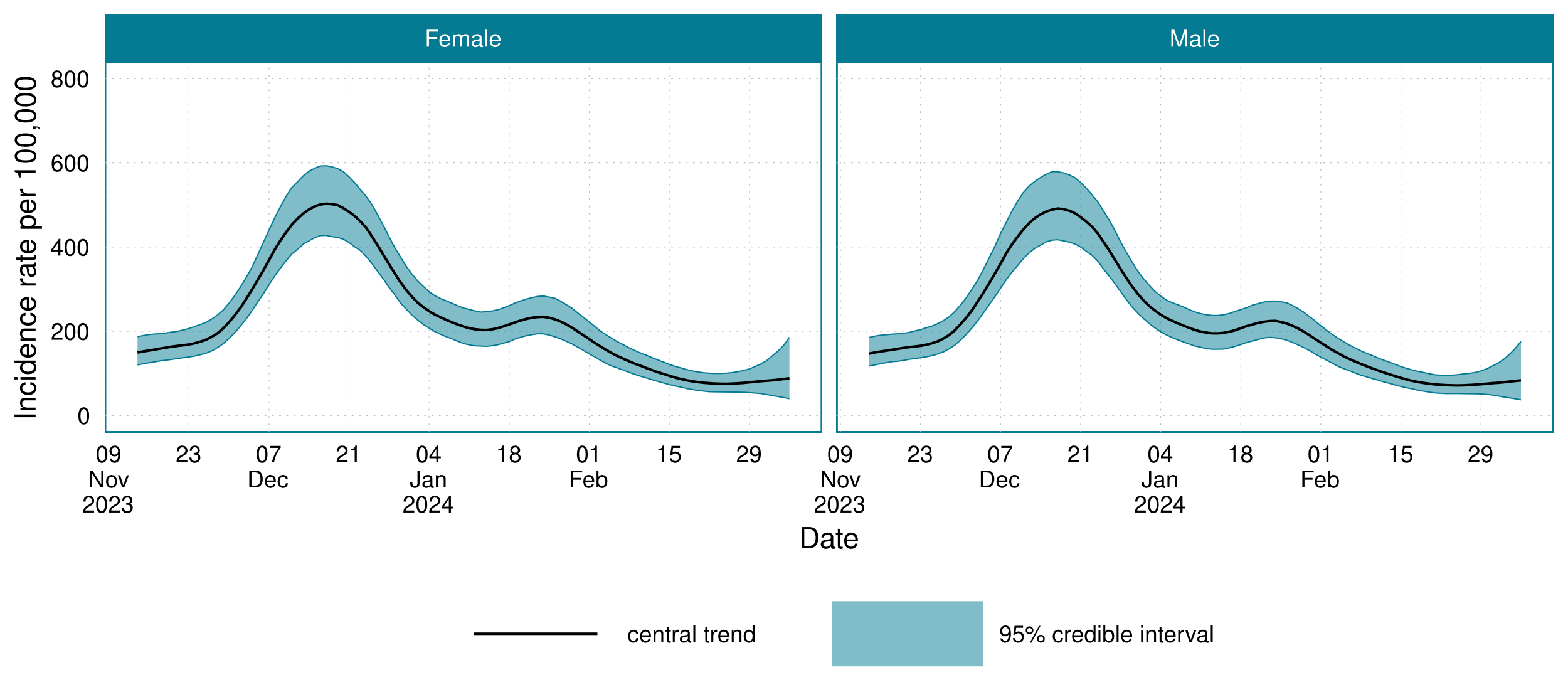

Supplementary Figure 6: The sex-stratified estimates of the incidence of SARS-CoV-2 infections for 14 November 2023 to 7 March 2024.

#### Average infections per individual

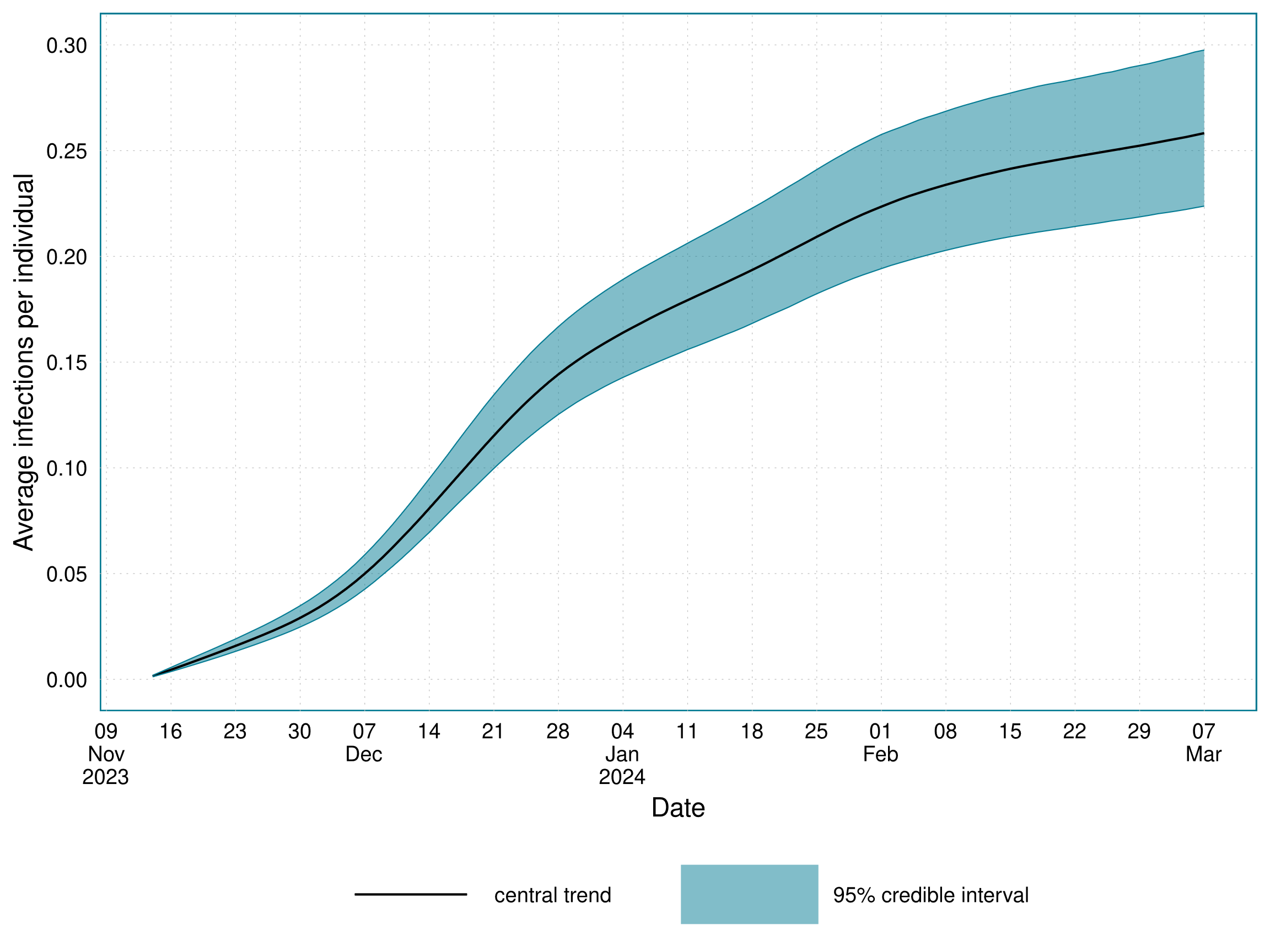
 Supplementary Figure 7: The time-varying average number of infections that had occurred per individual over the study period, 14^th^ November 2023 to 7^th^ March 2024, for England and Scotland.

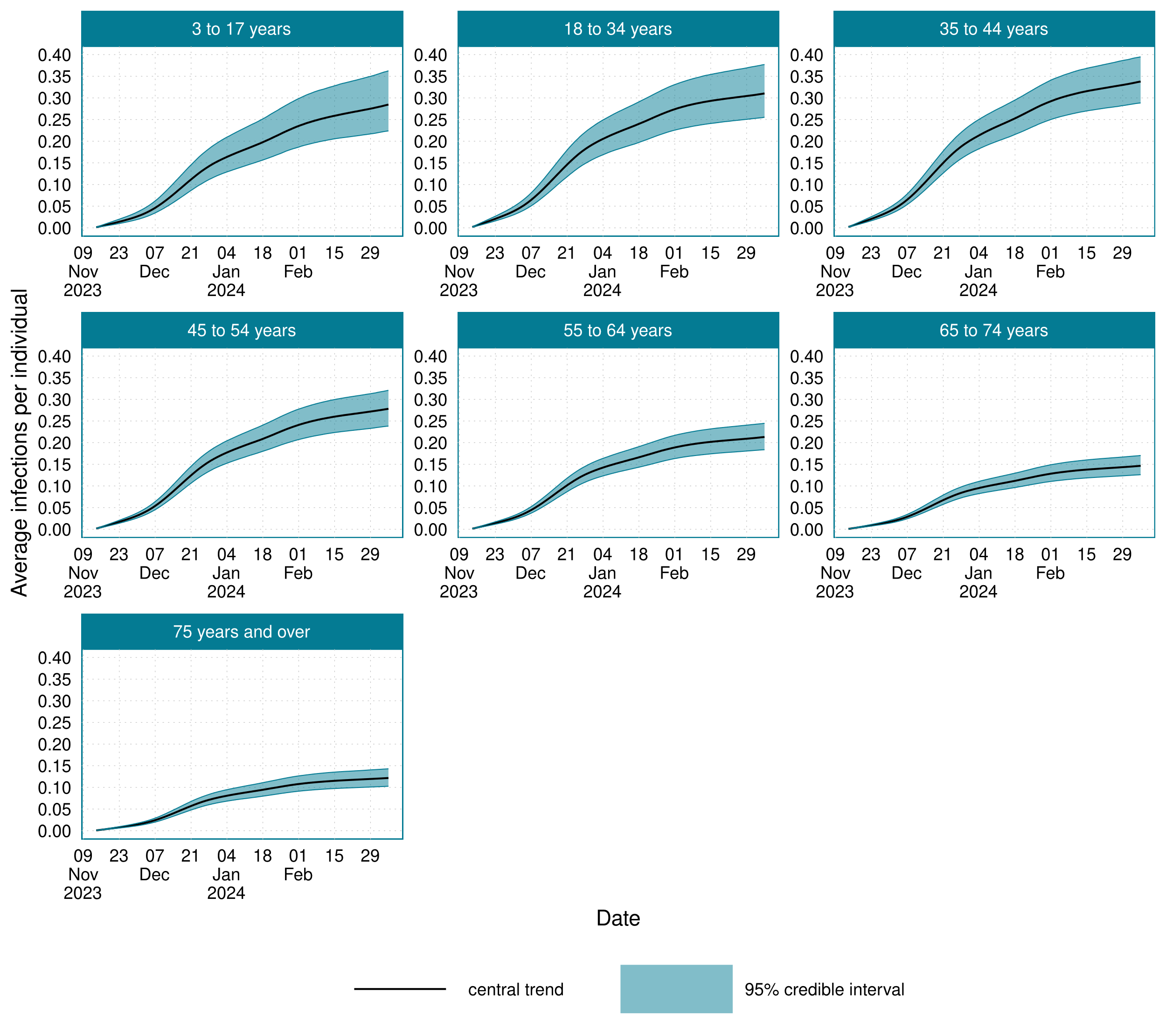

Supplementary Figure 8: The age-stratified average number of infections that had occurred per individual over the study period, 14^th^ November 2023 to 7^th^ March 2024, for England and Scotland.

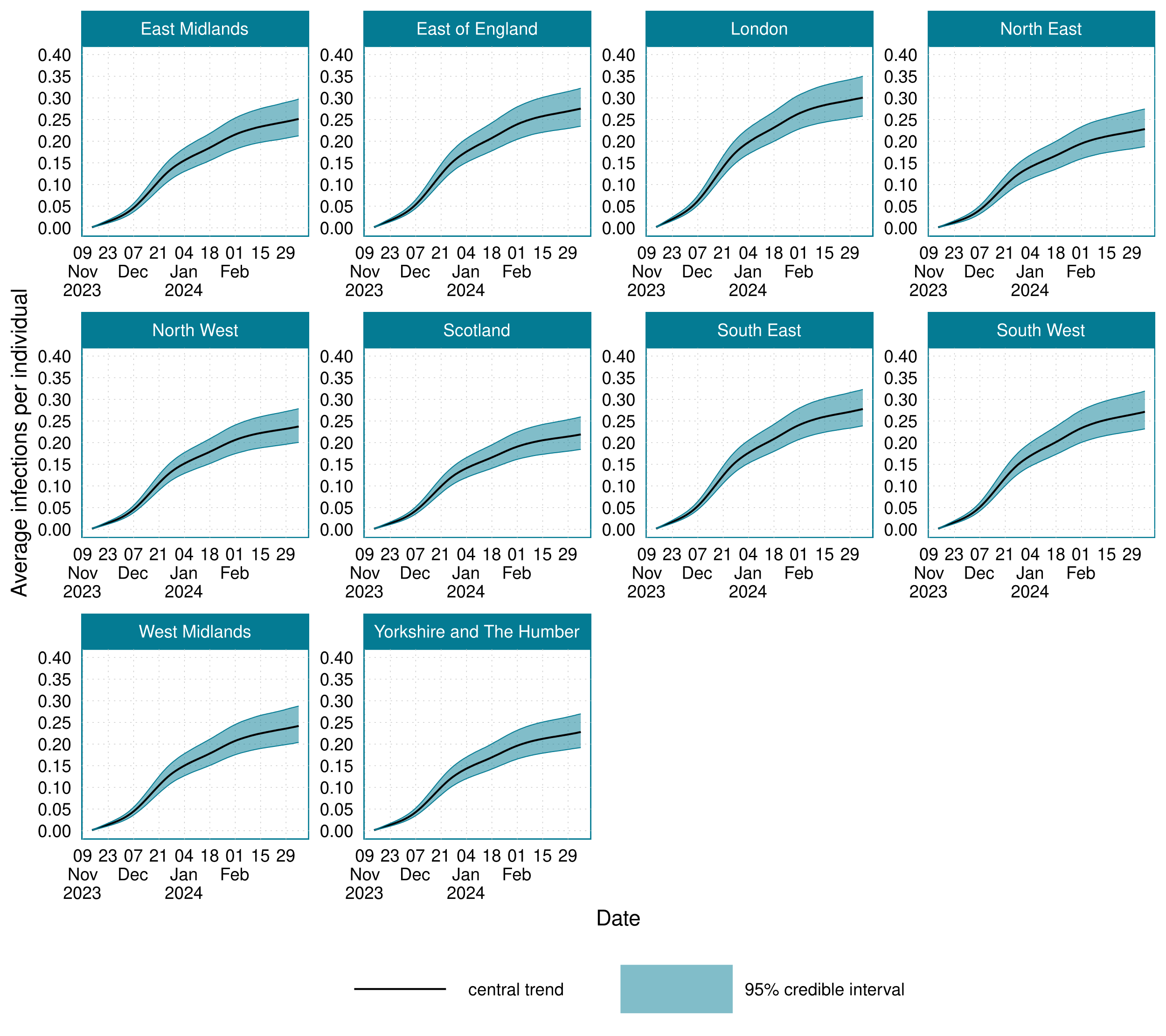

Supplementary Figure 9: The location-stratified average number of infections that had occurred per individual over the study period, 14^th^ November 2023 to 7^th^ March 2024, for England and Scotland..

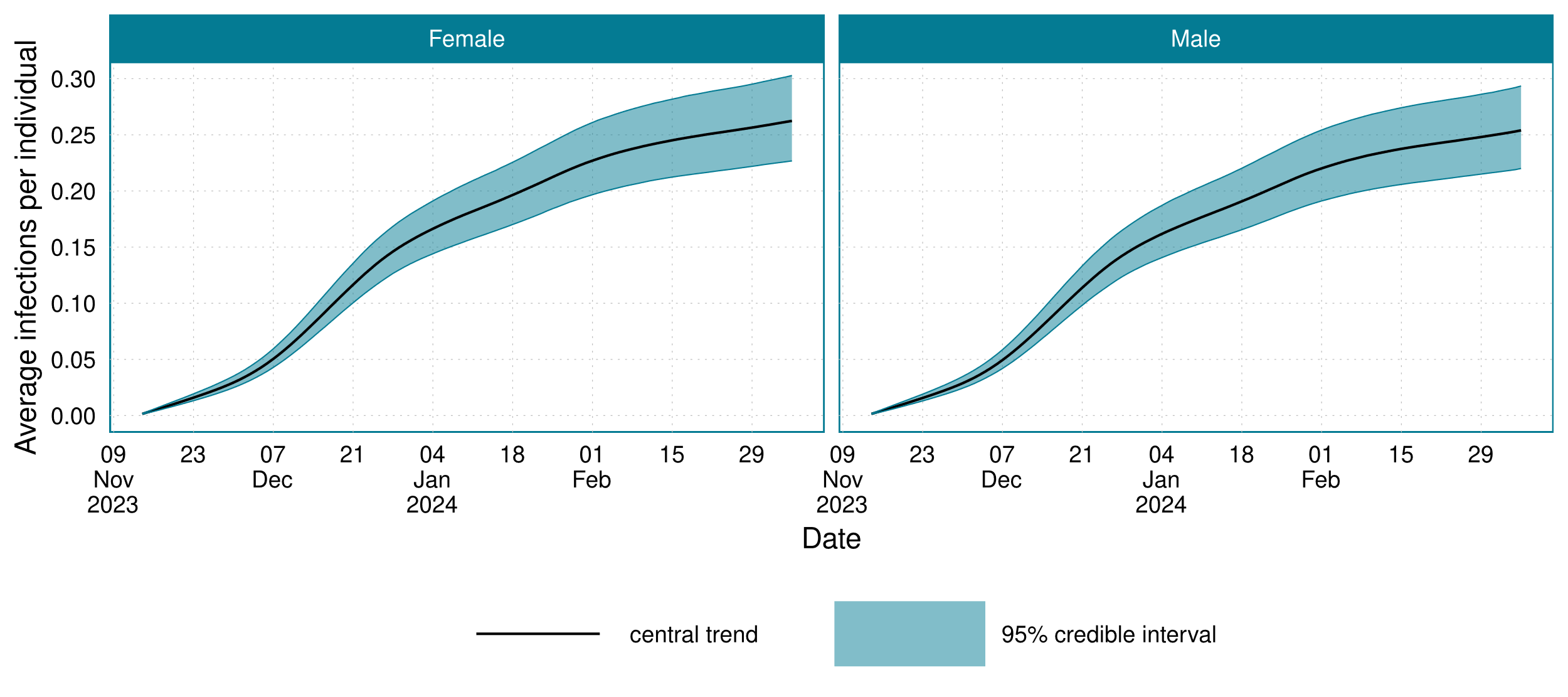

Supplementary Figure 10: The sex-stratified average number of infections that occurred per individual over the study period, 14^th^ November 2023 to 7^th^ March 2024, for England and Scotland.

#### Sensitivity

| Age group | Average sensitivity (%) | Minimum sensitivity (%) | Maximum sensitivity (%) | Maximum sensitivity date | Minimum sensitivity date |
| --- | --- | --- | --- | --- | --- |
| 3 to 17 years | 62.0 (95% CI: 55.8 to 67.9) | 57.5 (95% CI: 50.8 to 63.9) | 68.3 (95% CI: 62.3 to 73.9) | 08/12/2023 | 03/01/2024 |
| 18 to 34 years | 73.5 (95% CI: 68.8 to 77.8) | 70.1 (95% CI: 64.8 to 74.9) | 78.3 (95% CI: 74.2 to 82.0) | 09/12/2023 | 05/01/2024 |
| 35 to 44 years | 74.3 (95% CI: 71.6 to 76.9) | 70.8 (95% CI: 67.6 to 73.9) | 79.2 (95% CI: 76.8 to 81.7) | 09/12/2023 | 05/01/2024 |
| 45 to 54 years | 72.3 (95% CI: 70.2 to 74.5) | 68.4 (95% CI: 65.7 to 71.0) | 77.7 (95% CI: 75.5 to 79.9) | 09/12/2023 | 04/01/2024 |
| 55 to 64 years | 74.1 (95% CI: 72.3 to 75.8) | 70.6 (95% CI: 68.4 to 72.7) | 79.0 (95% CI: 77.2 to 80.9) | 09/12/2023 | 04/01/2024 |
| 65 to 74 years | 75.3 (95% CI: 73.6 to 77.0) | 72.1 (95% CI: 70.1 to 74.0) | 79.8 (95% CI: 78.1 to 81.8) | 09/12/2023 | 04/01/2024 |
| 75 years and over | 79.2 (95% CI: 76.9 to 81.3) | 76.2 (95% CI: 73.7 to 78.6) | 83.4 (95% CI: 81.3 to 85.4) | 10/12/2023 | 05/01/2024 |

Supplementary Table 6: Posterior summary statistics for the age-stratified population average test sensitivity values over time

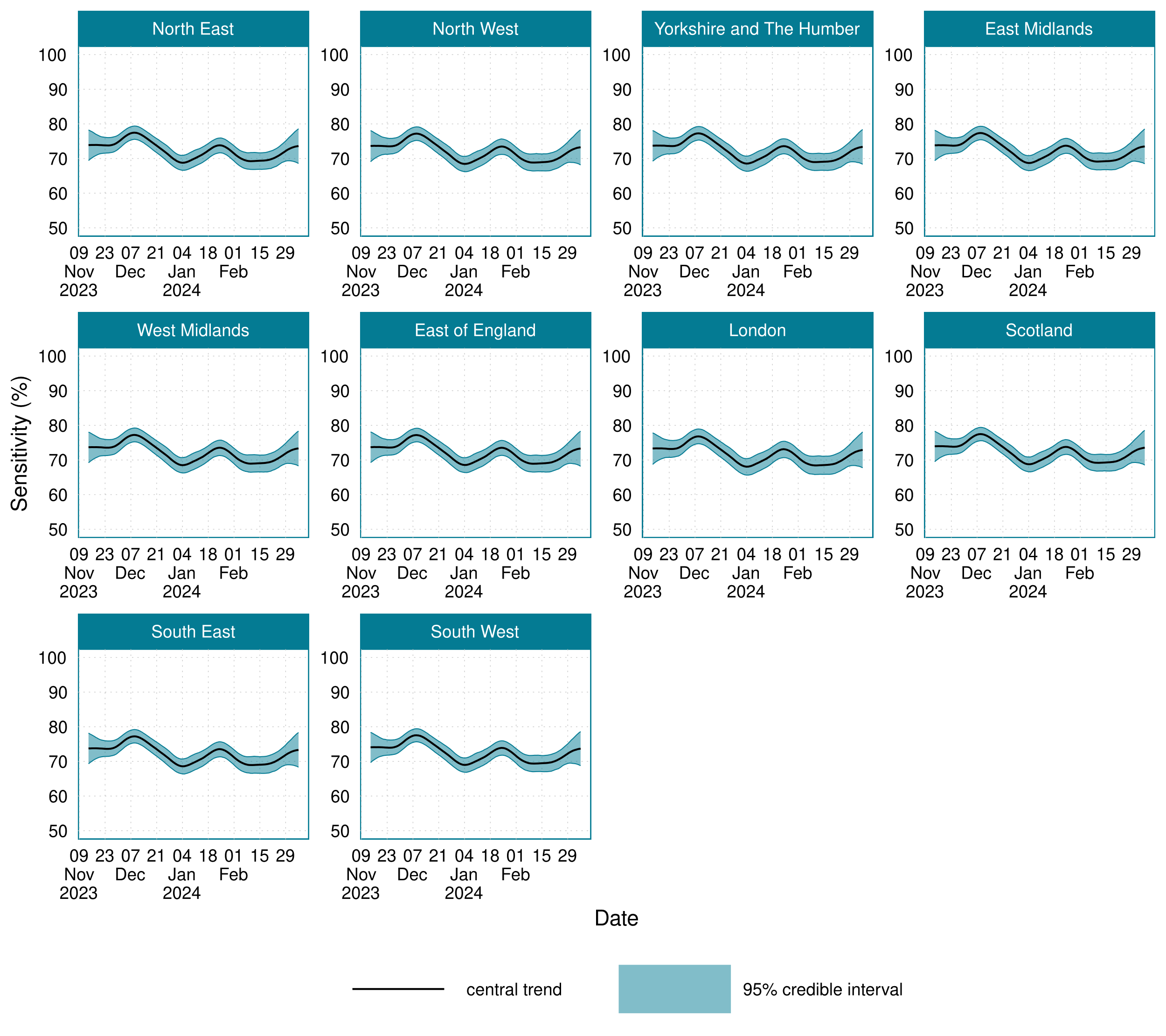

*Supplementary Figure 11: Location-stratified estimates of LFD sensitivity to SARS-CoV-2 infections for 14 November 2023 to 7 March 2024.*

| Location | Average sensitivity (%) | Maximum sensitivity (%) | Minimum sensitivity (%) | Maximum sensitivity date | Minimum sensitivity date |
| --- | --- | --- | --- | --- | --- |
| East Midlands | 72.3 (95% CI: 70.5 to 74.1) | 77.4 (95% CI: 75.4 to 79.3) | 68.7 (95% CI: 66.5 to 70.8) | 09/12/2023 | 04/01/2024 |
| East of England | 72.1 (95% CI: 70.3 to 73.9) | 77.2 (95% CI: 75.3 to 79.2) | 68.6 (95% CI: 66.4 to 70.7) | 09/12/2023 | 04/01/2024 |
| London | 71.7 (95% CI: 69.6 to 73.7) | 76.8 (95% CI: 74.7 to 78.9) | 68.1 (95% CI: 65.6 to 70.4) | 09/12/2023 | 04/01/2024 |
| North East | 72.4 (95% CI: 70.6 to 74.2) | 77.5 (95% CI: 75.6 to 79.4) | 68.8 (95% CI: 66.6 to 70.9) | 09/12/2023 | 04/01/2024 |
| North West | 72.1 (95% CI: 70.2 to 73.9) | 77.2 (95% CI: 75.2 to 79.1) | 68.5 (95% CI: 66.2 to 70.6) | 09/12/2023 | 04/01/2024 |
| Scotland | 72.4 (95% CI: 70.6 to 74.2) | 77.5 (95% CI: 75.6 to 79.4) | 68.8 (95% CI: 66.6 to 70.9) | 09/12/2023 | 04/01/2024 |
| South East | 72.2 (95% CI: 70.4 to 74.0) | 77.2 (95% CI: 75.3 to 79.2) | 68.6 (95% CI: 66.4 to 70.7) | 09/12/2023 | 04/01/2024 |
| South West | 72.5 (95% CI: 70.8 to 74.3) | 77.5 (95% CI: 75.7 to 79.4) | 69.0 (95% CI: 66.9 to 71.0) | 09/12/2023 | 04/01/2024 |
| West Midlands | 72.1 (95% CI: 70.3 to 74.0) | 77.2 (95% CI: 75.2 to 79.2) | 68.5 (95% CI: 66.3 to 70.7) | 09/12/2023 | 04/01/2024 |
| Yorkshire and The Humber | 72.2 (95% CI: 70.4 to 74.0) | 77.3 (95% CI: 75.3 to 79.2) | 68.6 (95% CI: 66.3 to 70.7) | 09/12/2023 | 04/01/2024 |

Supplementary Table 7: Posterior summary statistics for the location-stratified population average test sensitivity values over time
